# Evaluating the impact of an enhanced support implementation of the PReCePT (PRevention of Cerebral palsy in Pre-Term labour) quality improvement toolkit to increase the uptake of magnesium sulphate in pre-term deliveries for the prevention of neurodisabilities: study protocol for a cluster randomized controlled trial

**DOI:** 10.1101/2020.09.10.20190322

**Authors:** HB Edwards, MT Redaniel, BC Opmeer, TJ Peters, R Margelyte, Rejon C Sillero, W Hollingworth, P Craggs, EM Hill, S Redwood, JL Donovan, K Luyt

## Abstract

The UK’s National Institute for Health and Care Excellence (NICE) Preterm labour and birth guideline recommends use of magnesium sulphate (MgSO4) in deliveries below 30 weeks’ gestation to prevent cerebral palsy and other neurological problems associated with preterm delivery. Despite national guidance, the uptake of MgSO4 administration in eligible women has been slow. NHS England has rolled out the PReCePT Quality Improvement (QI) toolkit to increase uptake of MgSO4 in preterm deliveries. The toolkit is designed to increase maternity staff knowledge about MgSO4 and provides training and practical tools to help staff consider use in eligible women. The PReCePT trial will evaluate the effectiveness of an enhanced support model of implementing the QI toolkit, compared with the standard support model. The standard support arm (control) receives the QI toolkit and regional-level support for a midwife/obstetric ‘champion’. The enhanced support arm (intervention) receives this plus additional clinical backfill funding and unit-level QI micro-coaching.

This is a cluster randomised controlled trial designed to include 48 maternity units randomised (2:1 ratio) to standard or enhanced support. Units are eligible for inclusion if they have ten or more pre-term (< 30 weeks’ gestation) deliveries annually and MgSO4 uptake of 70% or less. Randomisation is stratified by previous level of MgSO4 uptake. The QI intervention is implemented over nine months. All units are followed up for a further nine months. Blinding is not possible due to the nature of the intervention.

The primary outcome is the proportion of MgSO4 uptake amongst eligible women at follow-up, adjusting for uptake before implementation of the toolkit. The effectiveness of the intervention will be assessed using weighted linear regression on data from the National Neonatal Research Database. Semi-structured qualitative staff interviews will inform understanding of the process and outcomes. Economic evaluation will describe total costs and cost-effectiveness.

**STRENGTHS AND LIMITATIONS:** *Strengths:*

- The first randomised controlled trial comparing two models of supporting the implementation of a Quality Improvement toolkit in perinatal medicine.
- A comprehensive evaluation, involving quantitative, qualitative and process measures including costs, to assess impact of the toolkit on the uptake of magnesium sulphate and team working.
- The National Neonatal Audit Programme (NNAP) and National Neonatal Registry Database (NNRD) provides robust routine data collection infrastructure for the primary outcome, also allowing future assessment of sustainability within participating units as well as uptake across the country.

*Limitations:*

- This pragmatic trial will reflect the conduct of scaling up a local initiative to a national level, where adherence to trial timelines may vary due to differences in local settings, procedures for permissions/approvals, and team capacity.
- Communication about the trial through formal and informal media channels may raise general awareness and thus improve background uptake nationally. Such contamination across trial groups may require assessment and adjustment in sensitivity analyses.

## INTRODUCTION

Preterm birth is the leading cause of neonatal mortality and morbidity,^1^ and specifically brain injury and cerebral palsy (CP).^2–4^ Around 1% of births in developed countries are very preterm (less than 30 weeks’ gestational age (GA)).^5^ While around 90% of very preterm infants survive beyond the postpartum period,^6^ it is estimated that approximately a third develop neurodisabilities including CP, blindness, deafness, and cognitive impairment.^7–9^ Around 10% of very preterm births in developed countries result in CP.^3,6,10^

Antenatal magnesium sulphate (MgSO4) therapy given to women at risk of preterm birth reduces the risk of CP in their child by around 30% (relative risk 0.68; 95% confidence interval 0.54 to 0.87).^11^ At under 30 weeks’ gestation, the number needed to treat to prevent one case of CP is 37 (95% CI 23 to 102).^12^ CP has a significant burden both for individuals and families^13^ and healthcare services, with an estimated lifetime cost per person (including health care, productivity, and social costs) of €830,000.^14,15^ Approximately 1,400 cases of brain injury among preterm babies could potentially be avoided by consistent administration of MgSO4 during labour each year in the UK, including 200 cases of CP annually in England.^12^

Since 2015 the UK National Institute for Health and Care Excellence (NICE) has recommended administration of MgSO4 in very preterm deliveries as a core part of maternity care.^16^ Failure to comply with this guideline is considered sub-optimal care. Uptake of MgSO4 in eligible women in the UK has historically been low compared with the rest of the developed world.^17,18^ For infants below 30 weeks’ gestation, the UK National Neonatal Audit reported that in 2017, only 64% of eligible women received MgSO4.^19^ There is high variation in uptake between different regional networks (range 49% to 78%).^19^ While there is evidence that uptake has been increasing (from 9% reported in 2012),^20^ many eligible women are still not receiving this important intervention.

The PReCePT Quality Improvement (QI) toolkit was developed to increase knowledge and awareness among maternity unit staff about MgSO4 as a neuroprotective agent in preterm deliveries.^21^ It provided practical tools and training to help staff consider MgSO4 in eligible women. It was co-designed by clinical teams and mothers who had experienced preterm birth. The PReCePT pilot study, set in five maternity units in the West of England, increased the MgSO4 uptake from an average baseline of 21% over the two years preceding the project to 88% by the end of the project.^21^ Improvements were observed for all participating units, although rates of uptake varied between maternity units.^21^

Based on the success of the PReCePT pilot, NHS England funded a national roll out of the intervention (National PReCePT Programme (NPP)). The NPP aims to support all maternity units in England to increase their use of MgSO4 to 85% of eligible women by 2020.The NPP was rolled out by the regional Academic Health Science Networks (AHSNs), whose role is to facilitate health innovations to improve health outcomes.

### Trial justification

The PReCePT pilot demonstrated that a QI package with bespoke unit-level coaching and backfill was effective in improving MgSO4 uptake. The National PReCePT Programme uses a reduced version of this package, more focused on providing resources for self-engagement. It is not clear if this reduced level of support will be sufficient to improve MgSO4 uptake to the target level. This trial compares the standard support as used in the NPP, with the enhanced support model as used in the original PReCePT pilot.

### Objective

The PReCePT trial described in this protocol paper is designed to compare the effectiveness, cost-effectiveness and sustainability of the enhanced support model compared with the standard level of support in encouraging increased use of MgSO4 amongst eligible women. Comparative evidence between the two adoption models will inform the method of optimal future UK spread.

## METHODS

### Trial Design

This is an open cluster randomised controlled trial set in NHS England maternity units. Each maternity unit is a “cluster”. The two trial arms (allocation ratio 2:1 control to intervention) are:

Control group (standard support): implementation of the PReCePT QI toolkit as guided by the NPP and regional AHSN. This includes provision of PReCePT QI materials (pre-term labour proforma, staff training presentations, parent leaflet, posters for the unit, learning log^22^), regional level QI training and support, and up to 90 hours funded backfill per unit for the midwife champion. Implementation is led by local midwives and an obstetrician champion, selected internally by each unit. (Table 1)

**Table 1:** Trial groups

|  | **Control (group 1, standard support)** | **Intervention (group 2, enhanced support)** |
| --- | --- | --- |
| PReCePT QI toolkit | Clinical guidance;<br>Pre-term labour proforma template;<br>Staff training presentations;<br>Parent leaflet;<br>Posters for display on the unit to raise staff awareness;<br>A QI Learning Log;<br>Project Dashboard;<br>Pens, magnets, lanyards and other aide-mémoires to promote MgSO<sub>4</sub> to unit staff (if purchased) | As per standard support group |
| QI training | Regional level QI training and guidance to adapt materials for local use, cascaded from AHSN | As per standard support group |
| Regional support | Support from a regional level neonatal lead and AHSN lead | As per standard support group |
| Local obstetrician champion | Local obstetrician identified by unit to guide and oversee local implementation | As per standard support group (named as joint PI, at discretion of local site) |
| Funded time for local midwife champion | Funded time of up to 90 hours per unit (on average 2 hours per week) | As per standard support, PLUS funding for up to 90 extra hours backfill, on average over 12 months, to enable the midwife to embed the QI toolkit within their team |
| Funded time for local neonatologist champion | None | Funded time for a local neonatologist PI, working on average 0.5 PA (2 hours) per week over 12 months, to provide clinical leadership in local unit (fixed term contract or secondment from an NHS organisation)<br>0.5 PA backfill may be split with obstetrician PI, at discretion of local site |
| QI coaching | None | Structured coaching in local unit from an experienced QI coach. To include:<br>First visit where the QI coach will work with local unit to create a bespoke implementation plan;<br>Telephone coaching in liaison with the local champion(s), with occasional face to face visits as logistics permit;<br>Ongoing dedicated support to help embed the QI toolkit within local unit;<br>Final visit to support local unit to tie up data collection and plan for ongoing sustainability |
| Learning events | None | Funding for up to three members of staff from local unit to attend three learning events. These bespoke learning events will be held every 2-3 months during the period of implementation and will bring together teams from other Group 2 units to share their activity and learning on how they are implementing the PReCePT QI toolkit and working to address issues and challenges |
| Celebration event | None | Provision of an android tablet to be used by the local midwife champion to micro-coach colleagues, plus a small fund for purchasing study collateral (pens, magnets, lanyards, aide-mémoires), if required |
| Collateral funding | None | Funding for up to three members of staff from local unit to attend a celebration event which will bring together teams from all Group 2 units to wrap up the study and to share experiences, learning and success. |

Intervention group (enhanced support): implementation of the PReCePT QI toolkit as for the standard support group, plus individual unit-level coaching by an experienced QI coach (a first in-person visit, a final in-person visit, and regular telephone coaching during the nine months implementation phase), a computer tablet for micro-coaching staff, access to learning and celebration events, an additional 90 hours backfill funding for the local midwife champion, and 0.5PA per week of funded backfill for the local neonatologist champion. At each unit’s discretion this 0.5PA backfill can be shared between the neonatologist and obstetrician champion. (Table 1)

The trial randomisation and implementation is aligned with the NPP timeframe as the trial is embedded within the NPP. The NPP is implementing the PReCePT QI toolkit in two waves, starting in May and September 2018. This staggered approach is to accommodate differences in readiness of units to put logistical arrangements in place. The trial is aligned with these waves to maximize comparability between groups. The enhanced QI support will be implemented in the intervention units for nine months after randomisation (December 2018 to August 2019 for first wave units; January 2019 to September 2019 for second wave units). The trial units will have a nine-month follow-up period after the end of the implementation phase. (Figure 1)

### Eligibility criteria

Maternity units in England participating in the NPP with ten or more pre-term (< 30 weeks gestation) deliveries annually and with MgSO4 uptake of 70% or less. Eligibility criteria are assessed from National Neonatal Audit Programme (NNAP) data from 2017. Units that took part in the PReCePT pilot are excluded.

### Consent

Written informed unit-level consent is required for participation. The clinical service lead for maternity and neonatal care at each eligible maternity unit is sent an invitation letter, unit information sheet describing the project, and consent form. On the advice of the UK National Health Service (NHS) Health Research Authority (HRA), consent was not obtained from individual women. This is because at the patient-level, only anonymous routinely collected data is used, and clinical guidance on the appropriate care for each individual woman is unaffected by either trial arm, or even whether or not their hospital is taking part in the study. For qualitative interviews with individual unit staff, individual consent will be obtained.

### Withdrawal Criteria

Units in the enhanced support model arm can withdraw at any time. They will then revert to the standard support model and be followed up accordingly. Their data will be collected and included as planned and analysed according to trial allocation (intention to treat). An exit interview will be requested to assess reasons for withdrawal. Staff participating in interviews can withdraw at any time and if they do their data will not be used in analysis.

### Sample size

The sample size of the enhanced support group is limited to 16 maternity units to fit within the trial budget. Based on results from the PReCePT pilot study^21^ and 2016 NNAP data, we anticipate MgSO4 uptake of approximately 38% and 80% in the two trial arms. With a 2-sided 5% significance level this study will have 86% power to detect an absolute difference of 40 percentage points in MgSO4 uptake at follow-up between the control and intervention groups (based on a 2:1 randomisation ratio). As the planned analysis is at the cluster(maternity unit) level, this removes any clustering effects that could impact on sample size calculations. NNRD data reports that during 2017, the target 48 maternity units had a mean of 30 preterm births (interquartile range 14 to 41).

### Randomisation

Maternity units are the units of randomisation. Of the eligible and consenting units, 48 are planned to be allocated within the trial at a 2:1 ratio (that is, 32 control and 16 intervention). Randomisation will occur in two waves in line with the NPP’s phased approach of starting the programme in two waves. In both wave one and wave two, 16 units are planned to be allocated to the standard support model arm, and eight units to the enhanced support model arm.

To reduce imbalance between groups, units will be stratified by 2017 MgSO4 uptake rates. Stratification groups based on consenting units are 0-39.9%, 40-49.9%, 50-59.9%, and 60-70.9%. For each trial arm, four reserve units will be selected and included in the randomisation, in case of unit drop-out.

Randomisation will be performed with Stata package command *stratarand* and carried out by a statistician independent of the trial and the NPP.

Due to the nature of the interventions it is not possible to conceal the allocation to members of the research team and hospital staff.

#### Outcomes

The primary outcome for the trial is the unit-level uptake of MgSO4 administration among eligible women (preterm birth < 30 weeks’ gestation) defined as whether or not the mother received MgSO4 prior to delivery. This is measured at the end of the trial and will be expressed as the percentage of eligible mothers receiving MgSO4 amongst all eligible mothers. To enable comparison with national reported data, we will be using the 2017 NNAP method of omitting mothers with missing/not available MgS04 data from both the numerator and denominator. We will conduct a sensitivity analysis to assess whether there is selection bias associated with the exclusion of these mothers.

We will consider secondary outcomes to further evaluate effectiveness in other respects, as well as investigations into the process of implementation, and an economic evaluation. For effectiveness we will additionally evaluate: trend in uptake (testing for step-change / change in trend) before, during and after implementation; longer-term trends in uptake over 2011-2019; reasons MgSO4 was not given in eligible women; whether the impact of the QI toolkit is affected when adjusting for potential confounding factors; whether the intervention was carried out as intended; staff experience; and data quality.

To evaluate the process of implementation at each unit we will explore: proportion and type of staff receiving training; number of and time required for training sessions; number and size of staff meetings for feedback and discussion; extent of other ongoing research / QI projects and previous QI experience; adherence to the PReCePT QI toolkit; staff confidence, involvement and engagement; organisational factors such as restructuring, understaffing, changes in management; and professional or cultural issues that could affect implementation.

For the economic evaluation we explore time and resources required in both intervention and control groups, cost associated with backfill for local clinical champions, total cost associated with each support model, and incremental cost-effectiveness ratio.

#### Analyses

The trial will use multiple methods to evaluate the enhanced QI support compared with the standard support.

### Effectiveness data collection and evaluation

We will use anonymised patient level extracts of the UK National Neonatal Research Database (NNRD) from units participating in the trial^23^. Data on MgSO4 use is collected routinely in BadgerNet, the clinical audit database completed by clinicians in every neonatal unit in England. BadgerNet data is transferred quarterly to the NNRD. Fields relating to the MgSO4 care pathway are mandatory and are regarded as good quality (over 70% completeness) since 2015. Data in the NNRD undergoes multiple quality assurance procedures and is considered to have high accuracy and completeness.^23,24^

The TeamSTEPPS Teamwork Perceptions Questionnaire (T-TPQ)^25^ will be administered to all units in both trial arms at the start (months one to three) and end of the implementation period (month nine). This measures any change in levels of collaborative maternity and neonatal team functioning, leadership, support and communication. It will be completed by the three local champions at each unit (champion midwife, neonatologist and obstetrician) to get a range of perspectives on perinatal teamworking.

To compare the effectiveness of enhanced support versus the standard support model, we will be using weighted linear regression to model MgSO4 uptake at the end of follow-up, adjusted for baseline MgSO4 uptake. We will use a regression-based adjustment for baseline and will adjust for clustering by conducting the regression with the cluster (maternity unit) as the unit of analysis.^26^ Baseline MgSO4 uptake is the uptake reported by the unit in the 12 months prior to randomization. Post-intervention MgSO4 uptake is the uptake reported by the unit at the end of the trial.

Multilevel mixed-effects models will be used to adjust for the different maternity unit characteristics and the effects of the AHSN structure. Factors adjusted for will include: NPP wave (one or two), level of neonatal unit (secondary or tertiary), unit annual number of births, previous QI experience (all data collected via a baseline questionnaire). Levels of maternal hypertension, gestational age at delivery, and antenatal steroid administration (unit-level averages measured at baseline, data from the NNRD) will also be adjusted for. For multiple births, in order to remain consistent with NNAP reporting, we will only include data on one baby (the first born) from each multiple birth. For describing baby-level demographics we will include all babies from multiple births.

Multiple imputation using chained equations (MICE) will be used to impute missing variables using the “ice” command in Stata. Twenty datasets will be imputed with an imputation model including the outcome, exposure, and all covariables. We will examine possible impact of Missing Not At Random using sensitivity analysis.

For the intervention units only, QI coaches will also record monthly data on each units’ level of engagement and activity with PReCePT (both scored as at-risk, progressing, or on-track) and risks/issues encountered. This will be collected as part of their regular interaction with each unit to deliver coaching. Multivariable linear regression will be used to assess whether using the ‘ice’ command in Stata. Twenty datasets will be imputed with an imputation model these factors are associated with level of MgSO4 uptake in intervention group maternity units.

### Qualitative data collection and evaluation

To evaluate the implementation of the QI intervention in each unit (for example, level of compliance, whether it was delivered as intended, any local adaptations, any unexpected obstacles, the local context, and staff experience), semi-structured qualitative interviews will be conducted with staff. Interviews will either be face-to-face, by telephone or video-call. These will be recorded, transcribed and analysed using the framework method.^27^

Criterion based sampling (trial arm, number of births per year, baseline rate of MgSO4 uptake, recent Care Quality Commission (CQC) ratings on units’ leadership and patient safety performance) will be used to select up to 20 trial units. We will purposively sample two to three participants at each site in the roles of midwife, obstetrician and/or neonatologist.

Interviews will be analysed using the framework method.^27^ The matrix output of summarized data will allow analysis by case (site, professional group, individual) and by code (in relation to a particular theme such as intervention fidelity). This allows comparison of data across as well as within cases to inform understanding of outcomes.

### Economic data collection and evaluation

We will conduct a policy cost-effectiveness evaluation to compare the cost-effectiveness of the enhanced support model versus the standard support model.^28,29^

To measure resource use at each unit we will use information provided by the National PReCePT Programme and AHSNs, and data collected via electronic proformas issued monthly to each trial unit and completed by local champions. These will record time spent preparing reports, at events, at staff training sessions, number and type of staff involved, and time spent receiving QI coaching/support.

Costs are estimated by multiplying the volume of resources used (mainly staff time) by the price of each resource unit (unit cost). Costs, for example based on staff salary band, will be valued using national unit cost estimates, where available.^30^ Mean total implementation costs per unit will be estimated for both support models. We will categorise costs according to the different phases of the QI in which they occur. Specifically, developmental costs; organising costs; executing costs; and sustainability costs.

The Incremental Cost-Effectiveness Ratio will be calculated and shows the additional costs required to achieve one additional percentage point improvement of MgSO4 uptake. Univariate sensitivity analyses will be carried out to evaluate the impact of assumptions and unit cost estimates on the results. Previous economic analyses^31,32^ have estimated the long-term cost-effectiveness of MgSO4 administration in preterm births. If enhanced support results in increased uptake of MgSO4 administration we will use this evidence to estimate the long-term cost-effectiveness of enhanced support in terms of costs per quality adjusted life year (QALY) gained.

#### Data monitoring

As this is a QI project, data monitoring will largely be completed at local level. The local neonatologist champion will have responsibility for monitoring data completion in their unit. As part of the NPP, the NHS National Patient Safety Measurement Unit (PSMU) will create a national dashboard demonstrating the data from BadgerNet on MgSO4 administration. Local units will be able to produce monthly reports to monitor performance. The trial team will also be able to monitor data collection for trial units and address any data concerns. Any concerns will be reported to the Trial Steering Group.

#### Public and Patient Involvement (PPI)

PPI for the trial builds on the involvement work that took place in the PReCePT pilot study.^21^ This used a co-design and co-production approach including a partnership with BLISS, a support organisation for mothers experiencing pre-term births, and two mothers who had experienced pre-term births. The two mothers were part of the steering group for the project and were involved in trial design. People in Health West of England (PHWE), a shared regional public involvement resource based in the West of England, also helped to shape the design. A reference group of relevant stakeholders will help guide dissemination of findings.

#### Ethics and regulatory considerations

After discussion with the UK National Health Service (NHS) Health Research Authority, they gave authorisation that this trial does not require Research Ethics Committee (REC) approval as it is a low risk study involving NHS staff as participants. The trial was peer reviewed by an independent expert panel of reviewers as part of the funding application process. The panel was convened by the funder (The Health Foundation). The sponsor (University Hospitals Bristol and Weston NHS Foundation Trust) did not deem further peer review to be necessary for this low-risk research. We declare that the Chief Investigator and the committee members have no significant competing financial, professional, or personal interests that might have influenced the development of the trial design.

#### Other trial information

IRAS number 242419, ISRCTN 40938673, Trial Sponsor’s reference CH/2017/6417, Funder’s reference 557668. The Health Foundation funded this trial.

This research was supported by the National Institute for Health Research (NIHR) Applied Research Collaboration West (NIHR ARC West). The views expressed in this article are those of the author(s) and not necessarily those of the NIHR or the Department of Health and Social Care.

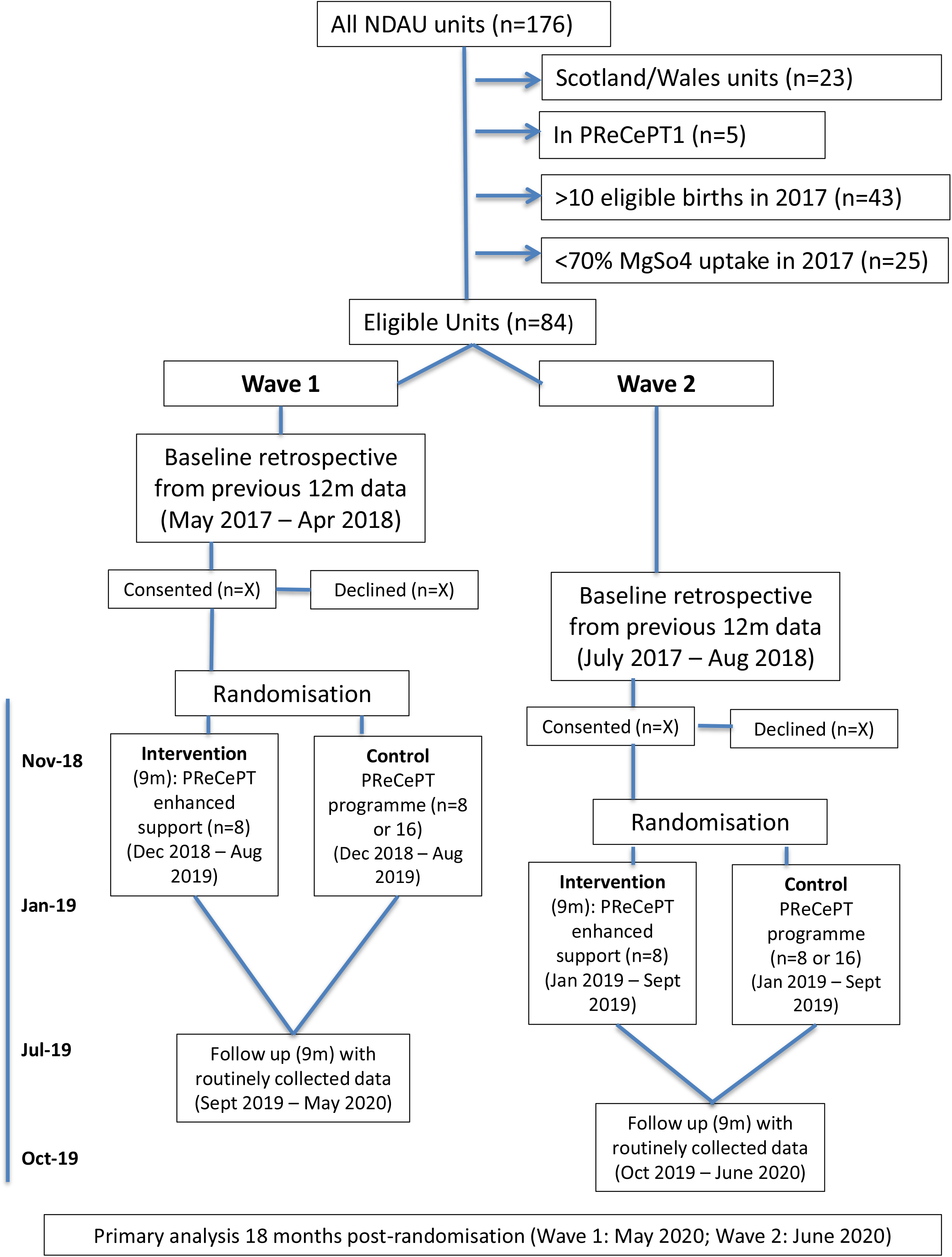

## Data Availability

All data relevant to the study are included in the article or uploaded as supplementary information. This article describes a trial protocol and as such, data from trial results are not yet available.

## Notes

### Competing Interest Statement

The authors have declared no competing interest.

### Clinical Trial

ISRCTN 40938673

### Funding Statement

The Health Foundation funded this trial. Funders reference 557668. This research was supported by the National Institute for Health Research (NIHR) Applied Research Collaboration West (NIHR ARC West).

### Author Declarations

UK National Health Service (NHS) Health Research Authority reference 242419

